# Approach-Avoidance Tendencies Moderate the Relationship Between Fear of Movement and Physical Activity in Osteoarthritis

**DOI:** 10.1101/2025.01.23.25321044

**Authors:** Miriam Goubran, Christian Zammar, Santiago Tellez Alvarez, Élodie Héran, Sara Proulx, Martin Bilodeau, Matthieu P. Boisgontier

## Abstract

**Research question:** Do psychological processes, such as explicit attitudes and approach-avoidance tendencies toward physical activity, mediate or moderate the relationship between fear of movement and usual physical activity levels in people with osteoarthritis?

**Methods:** An online observational study was conducted in 197 participants, including 68 with osteoarthritis. Arthritis, fear of movement, usual physical activity level, and explicit attitudes were assessed using questionnaires. Approach-avoidance tendencies, an indicator of automatic attitudes, was derived from reaction times in an approach-avoidance task.

**Results:** Results showed that higher fear of movement was associated with lower physical activity levels in participants with osteoarthritis. This association was moderated by approach-avoidance tendencies toward physical activity, with a significant effect only in participants with an automatic tendency to avoid physical activity or a weak tendency to approach it.

**Conclusion:** This study suggests that in adults with osteoarthritis, the detrimental effect of fear of movement on usual physical activity levels may be mitigated by strong automatic tendencies to approach physical activity. Since these tendencies result from the automatic activation of affective memories, health professionals should consider not only promoting physical activity but also ensuring its association with positive emotional experiences.

## 1. INTRODUCTION

Arthritis is an inflammatory disease that primarily affects synovial joints and is often associated with pain, stiffness, swelling, and reduced range of motion, which can ultimately lead to permanent disability.^1–3^ Exercise programs have been shown to be safe and effective in improving the condition of patients with arthritis, particularly in reducing pain and enhancing muscle strength.^4,5^ However, physical inactivity is almost twice as common in people with arthritis (31%) as in people without arthritis (17%).^6^ In addition, some comorbidities may further limit the ability to participate in physical activity.

One of these comorbidities is fear of movement, defined as an excessive, irrational, and debilitating fear of moving, resulting from a sense of vulnerability to pain, injury, or a medical condition.^7^ Fear of movement results from the repeated expression of negative affective responses in relation to physical activity.^8^ Here, we propose that this repetition underpins the formation of negative associations driving automatic processes that manifest as attitudes and triggering avoidance impulses to physical activity. The prevalence of fear of movement in patients with osteoarthritis is 58%.^9^ This high prevalence, combined with the association between higher fear of movement and lower physical activity reported in a recent meta-analysis (r = -.25; 95% confidence intervals [95CI] = -.39 to -.10),^10^ may explain why people with arthritis are less active than those without arthritis.^6^ However, the mechanisms underlying this association remain poorly understood.

Attitude is defined as a psychological tendency to evaluate a stimulus with some degree of favor or disfavor that operates on two levels: automatic and explicit.^11^ Automatic (formerly implicit)^12^ attitudes are traces of past experience that remain introspectively unidentified and mediate favorable or unfavorable evaluation of a behavior.^13^ This evaluation results in an automatic positive or negative inclination toward that behavior.^11–13^ For instance, when an individual perceives a cue related to bodily movement, this may automatically activate affective memories (positive or negative) associated with the concept of physical activity.^14^ This activation may generate an impulse, or motor readiness, that favors either the approach or avoidance of physical activity.^15,16^ In contrast, explicit attitudes are deliberate, reflective evaluations that individuals are consciously aware of, can articulate, and have some control over.^17^

Stronger automatic tendencies to approach images depicting physical activity^18^ and more positive explicit attitudes toward physical activity^19^ have been shown to be associated with higher levels of physical activity engagement. In addition, both approach-avoidance tendencies and have been explicit attitudes shown to be influenced by health conditions, such as apathy.^19^ Therefore, these two psychological constructs may affect the relationship between another health condition, fear of movement, and physical activity engagement.

The objective of this study was to examine the relationship between fear of movement, explicit attitudes toward physical activity, approach-avoidance tendencies toward physical activity, and usual physical activity levels in adults with arthritis. We hypothesized that (1) fear of movement would be higher in people with arthritis than in those without arthritis; (2) explicit attitudes and approach-avoidance tendencies would mediate or moderate the effect of fear of movement on physical activity levels (Figure 1).

**Figure 1.**
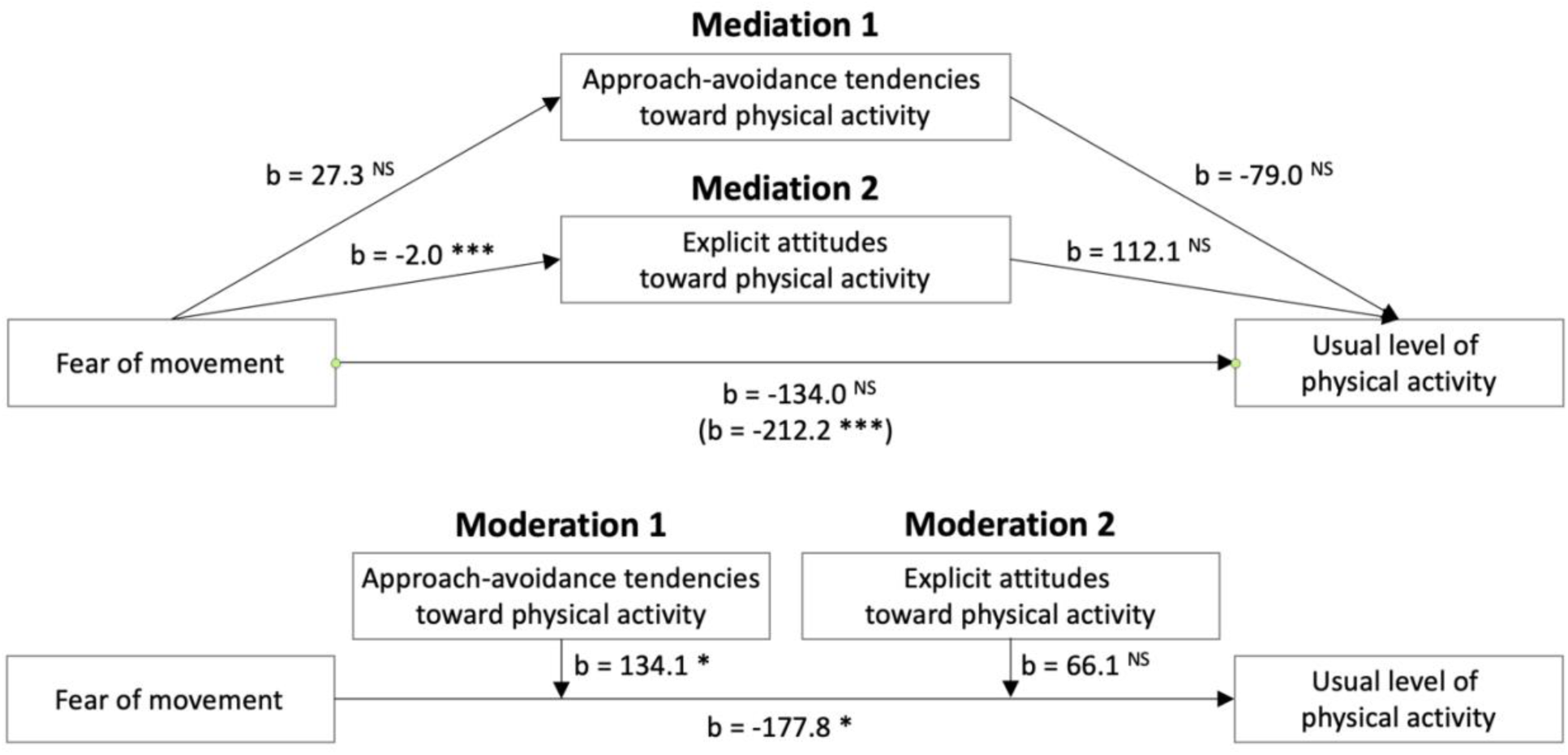
Tested mediation and moderation models in people with osteoarthritis. Notes: *p<.05; ***p<.001; NS = not significant; Notes: b = regression coefficient

## 2. METHODS

### 2.1. Population

Participants were recruited through social media (Facebook), posters at the Faculty of Health Sciences, University of Ottawa, and emails to associations of patients with arthritis and caregivers. Inclusion criteria were age 20–90 years and access to a personal computer, a laptop, or a tablet with internet. Informed consent was collected in accordance with the Declaration of Helsinki. The study was approved by University of Ottawa’s Research Ethics Boards (H-05-21-6791). All participants provided informed consent.

### 2.2. Power analysis

An a priori power analysis was conducted in G*power^20^ to estimate the minimum sample required for a two-tailed test with α = 0.05, power (1-β) = 90%, and a medium effect size f^2^ = 0.2.^21^ The analysis based on an F test in a linear multiple regression (R^2^ increase) that included two tested predictors and eight control variables estimated that a minimum total sample size of n = 67 was required for the moderation analysis. The sample size for the mediation analysis was lower as the models had one tested predictor per model and a maximum of 7 control variables.

### 2.3. Experimental protocol

#### 2.3.1. Procedures

Participants remotely and anonymously completed the study online using Inquisit software^22^ and provided information related to their arthritic condition, fear of movement, pain during exercise, usual level of moderate-to-vigorous physical activity, age, sex (male, female), gender (man, woman, non-binary, transgender man, transgender woman, other), weight, height, and explicit attitudes toward physical activity. One attention check question was included in the questionnaires: ‘Please answer ‘5’ to this question that allows us to verify that you actually read the questions.’ Automatic attitudes toward physical activity and sedentary behavior were tested using an approach-avoidance task.

#### 2.3.2. Self-reported variables

##### Arthritis

The presence of arthritis was derived from a question based on item PH006 of the Survey of Health, Ageing and Retirement in Europe.^23^ ‘Has a doctor ever told you that you had any of the following conditions?’ The participants who selected ‘arthritis, including osteoarthritis, or rheumatism’ but not ‘rheumatoid arthritis’ were considered as participants with osteoarthritis. The participants who selected both options (n = 12) or ‘rheumatoid arthritis’ only (n = 1) were not included in the analyses.

##### Chronic conditions

The total number of chronic conditions was used as a control variable in the analyses (Suppl. Material 1).

##### Fear of movement

Fear of movement was assessed using a modified version of the 11-item Tampa Scale of Kinesiophobia.^24^

##### Usual level of moderate-to-vigorous physical activity

The usual level of physical activity was derived from the short form of the International Physical Activity Questionnaire (IPAQ-SF), a self-administered questionnaire that identifies the frequency and duration of moderate and vigorous physical activity during the past seven days.^25^ The usual level of moderate-to-vigorous physical activity in minutes per week was used in the analyses.

##### Explicit attitudes

Explicit attitudes toward physical activity were assessed through two items based on two bipolar semantic differential adjectives on a 7-point scale (unpleasant-pleasant; unenjoyable-enjoyable). The statement begins with ‘For me, to participate in regular physical activity is …’.^18,26^ These two scores showed a very strong correlation (r = .939; p < 2.2×10^-16^), and their sum was used as an indicator of explicit attitudes in the statistical analyses.

##### Pain

Pain was measured to account for its potential confounding effect and was dervived from the statement: ‘During physical activity, I experience …’ with the possible responses ranging from 0 (‘no pain’) to 7 (‘pain as bad as it can possibly be’).

#### 2.3.3. Automatic approach-avoidance tendencies

Participants performed an approach-avoidance task (Figure 2A; Suppl. Material 1).^18,19,26^ Automatic tendencies were derived from reaction times (i.e., the time between stimulus appearance and key press) to approach or avoid stimuli depicting neutral, sedentary, and physical activity stimuli (Figure 2B).

**Figure 2.**
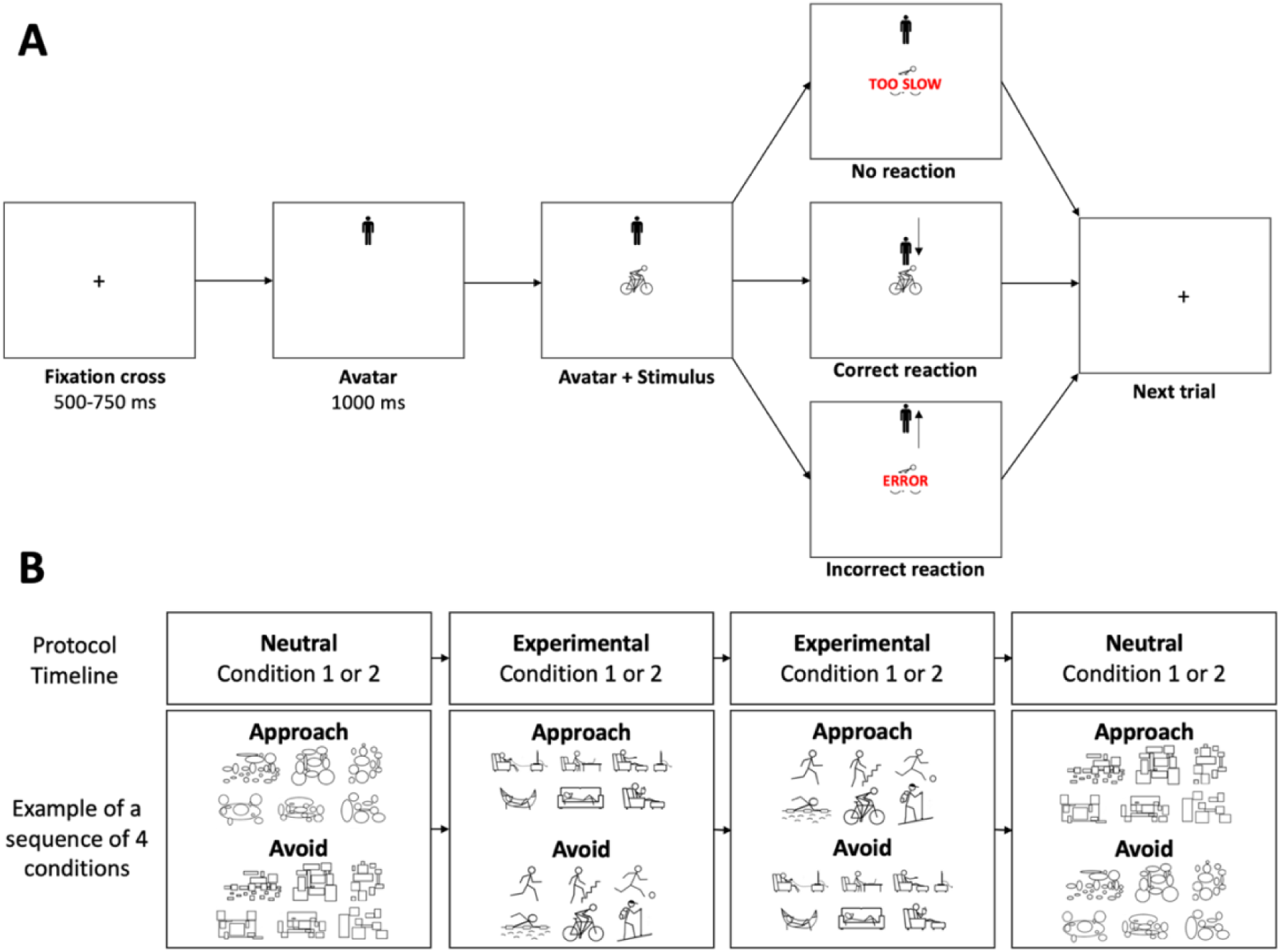
A. Illustration of a single trial of the approach-avoidance task in the condition where the participant is instructed to approach physical activity stimuli (and avoid sedentary stimuli – not shown). B. Timeline and stimuli of the approach-avoidance task. In Condition 1 (experimental and neutral), participants are instructed to move the avatar toward (approach) a specific type of stimuli (physical activity stimuli or rectangles) and away from (avoid) the other type of stimuli (sedentary behavior or ellipses, respectively). In Condition 2, the instructions are reversed: Participants move away from physical activity or rectangle stimuli and toward sedentary behavior or ellipse stimuli.

### 2.4. Statistical analyses

#### 2.4.1. Arthritis status and fear of movement

To test the effect of fear of movement on usual physical activity levels in people with and without arthritis, we performed multiple linear regression analyses testing the interaction between fear of movement and arthritis status (any arthritis vs. no arthritis) and ostseoarthritis status (osteorthritis vs. no arthritis), and including five control variables (age, sex, body mass index, pain, and number of chronic conditions). The lm() and confint() function from the ‘stats’ package in the R software environment^27^ were respectively used to conduct the models and compute 95CI. A separate analysis of participants with rheumatoid arthritis is reported in Suppl. Material 2.

#### 2.4.2. Mediation analysis

To examine the mediating effect of approach-avoidance tendencies and explicit attitudes toward physical activity on the association between fear of movement and usual physical activity levels in people with osteoarthritis only, we used the component approach, which involves three linear multiple regression models.^28^ Model 1 tests whether the independent variable (i.e., fear of movement) affects the outcome (i.e., physical activity). Model 2 tests the effect of the independent variable on the mediators (approach-avoidance tendencies [Mediation 1] or explicit attitudes [Mediation 2]). Model 3 tests both the independent variable and the mediators as simultaneous predictors of the outcome. Mediation is claimed if the above-mentioned effects are observed and if the ‘total effect’ of the dependent variable in Model 1 is larger in absolute value than its ‘residual effect’ in Model 3. Five control variables were included in these models (age, sex, body mass index, pain, and number of chronic conditions).

#### 2.4.3. Moderation analysis

To examine the moderating effect of approach-avoidance tendencies (Moderation 1) and explicit attitudes toward physical activity (Moderation 2) on the association between fear of movement and the usual level of physical activity engagement in people with osteoarthritis only, we examined the interaction effects of fear of movement with explicit attitudes and approach-avoidance tendencies toward physical activity on the dependent variable (usual level of physical activity). The control variables were the same as in the other models (age, sex, body mass index, pain, chronic conditions).

## 3. RESULTS

### 3.1. Descriptive results

Two hundred and fifty-six participants initiated the study. Fifty-six were excluded because they stopped the session before completing the study. Three participants who completed the full study were excluded because they answered the check question incorrectly. When participants reported height <50 cm or >250 cm or weight <30 kg or >250 kg, the data was removed and imputed by the mean value of the sample. The final sample of 197 participants included 116 participants without arthritis, 68 participants with osteoarthritis but no rheumatoid arthritis, and 13 participants with rheumatoid arthritis, including 12 participants who also had osteoarthritis (Table 1; Suppl. Figure 1). Participants with both osteoarthritis and rheumatoid arthritis were not included in the analyses. One male without arthritis identified themselves as a woman. All the other male (n = 75) and female participants (n = 121) identified themselves as men and women, respectively.

**Table 1.** Descriptive statistics stratified by arthritis status.

| Variable | No Arthritis<br>(n = 116) | Osteoarthritis<br>(n = 68) | Cohen's<br>d | p |  |
| --- | --- | --- | --- | --- | --- |
| Number of male / female participants | 53 / 63 | 20 / 48 |  |  |  |
| Fear of movement (mean ± SD) | 27.2 ± 11.6 | 31.7 ± 13.0 | 0.368 | .021 | * |
| Physical activity (mean ± SD) | 449.4 ± 537.2 | 425.9 ± 548.1 | -0.043 | .778 |  |
| Age (mean ± SD) | 55.3 ± 18.5 | 65.4 ± 11.3 | 0.621 | <2×10<sup>-16</sup> | *** |
| Pain (mean ± SD) | 2.64 ± 1.3 | 3.2 ± 1.5 | 0.427 | .009 | ** |
| Body mass index (mean ± SD) | 27.6 ± 8.8 | 27.4 ± 7.2 | -0.023 | .876 |  |
| Chronic Conditions (mean ± SD) | 1.1 ± 1.3 | 2.7 ± 1.3 | 1.178 | <2×10<sup>-16</sup> | *** |

Note: \*p<.05; \*\*p<.01; \*\*\*p<.001

### 3.2. Statistical results

#### 3.2.1. Effect of arthritis on the association between fear of movement and physical activity

The multiple linear regression showed a moderating effect of arthritis status (osteoarthritis vs. no osteoarthritis) on the association between fear of movement and the usual level of moderate-to-vigorous physical activity (b = 193.7; 95CI = -23.6 to 363.8; p = .026) (Suppl. Table 1). A simple effects analysis revealed that higher fear of movement was associated with lower physical activity in people with osteoarthritis (b = -150.4; 95CI = -287.5 to -13.2; p = .032), but showed no evidence in people without arthritis (b = 43.3; 95CI = -71.5 to 158.1; p = .457) (Figure 3).

**Figure 3.**
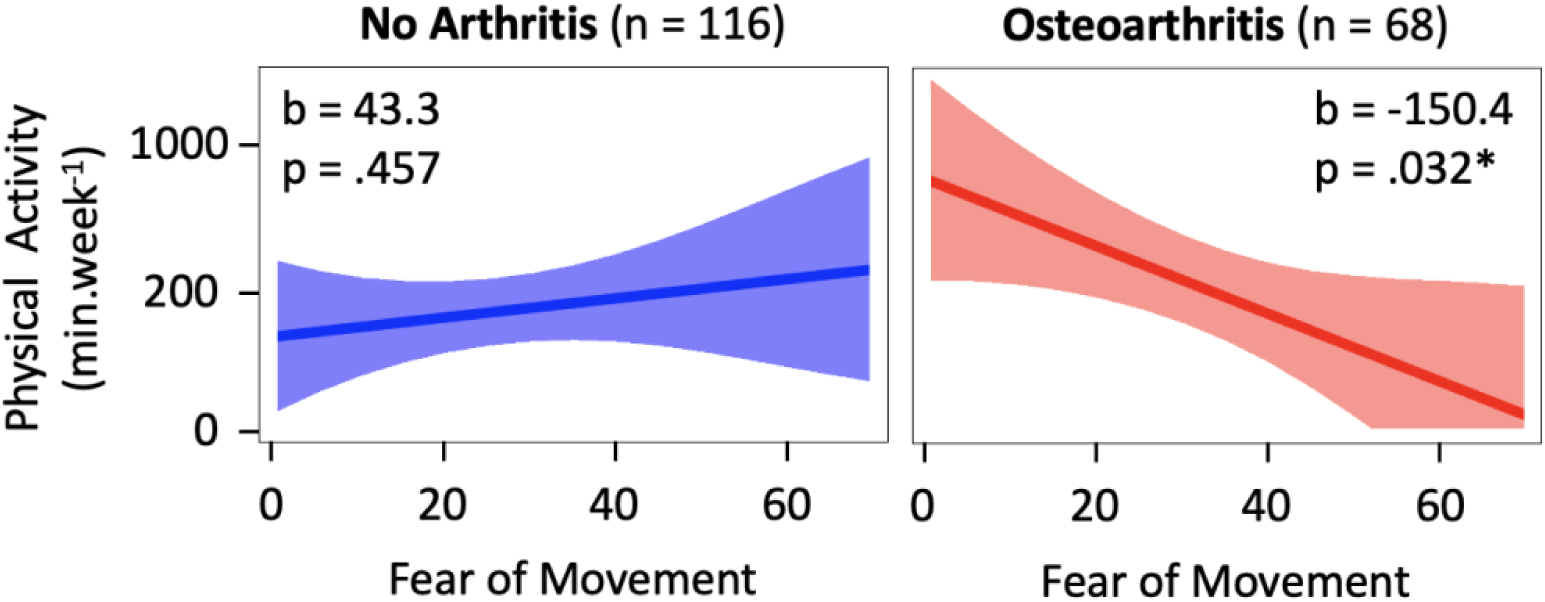
Simple effect of fear of movement on the usual physical activity level in participants without arthritis and with osteoarthritis. *p<.05; b = regression coefficients per standard deviation change in fear of movement; x axis is in raw scale to facilitate interpretation.

#### 3.2.2. Mediation analysis

In the sample of participants with osteoarthritis (n = 68), Model 1 of the mediation analysis showed an association between fear of movement and the usual physical activity levels (b = -212.2; 95CI = -360.7 to -63.6; p = 5.8 × 10^-3^). Model 2 for Mediation 1 showed no evidence of an association between fear of movement and approach-avoidance tendencies toward physical activity (b = 27.3; 95CI = -62.9 to 117.4; p = .547), ruling out these tendencies as a mediator. Model 2 for Mediation 2 showed an association between fear of movement and explicit attitudes toward physical activity (b = -2.0; 95CI = -2.9 to -1.2; p = 1.2 × 10^-5^). However, Model 3 showed no evidence of an association between explicit attitudes and the usual physical activity levels (b = 112.1; 95CI = -30.8 to 255.0; p = .134). Taken together, these results suggest that the relationship between fear of movement and usual physical activity levels is not mediated by approach-avoidance tendencies or explicit attitudes toward physical activity (Figure 1; Table 2).

**Table 2.** Mediation analysis: Estimated effects on the outcome measure (Model 1 and Model 3) and on the hypothesized mediators (Model 2A and 2B) in participants with osteoarthritis (n = 68)

| Variables | Model 1<br>Usual level of physical activity |  |  |  | Model 3<br>Usual level of physical activity |  |  |  |
| --- | --- | --- | --- | --- | --- | --- | --- | --- |
| | $b$ | 95CI | $p$ | | $b$ | 95CI | $p$ | |
| (Intercept) | 553.1 | 255.8;850.4 | $4.3 \times 10^{-4}$ | *** | 512.1 | 216.1;808.0 | .001 | ** |
| Age | -234.6 | -441.0;-28.1 | .027 | * | -240.8 | -446.3;-35.3 | .022 | * |
| Sex | 186.4 | -96.3;469.0 | .192 |  | 179.3 | -98.3;456.9 | .201 |  |
| Body mass index | 101.1 | -57.0;259.3 | .206 |  | 110.6 | -46.6;267.8 | .164 |  |
| Pain | 85.0 | -59.6;229.6 | .244 |  | 88.9 | -52.9;230.6 | .214 |  |
| Chronic conditions | -26.2 | -136.4;83.9 | .636 |  | -9.0 | -118.1;100.1 | .870 |  |
| Approach tendencies |  |  |  |  | -79.0 | -183.0;25.1 | .122 |  |
| Explicit attitudes |  |  |  |  | 112.1 | -30.8;255.0 | .134 |  |
| Fear of movement | -212.2 | -360.7;-63.6 | $5.8 \times 10^{-3}$ | *** | -134.0 | -305.0;808.0 | .547 | |

| Variables | Model 2 for Mediation 1<br>Approach-avoidance tendencies |  |  |  | Model 2B for Mediation 2<br>Explicit attitudes |  |  |  |
| --- | --- | --- | --- | --- | --- | --- | --- | --- |
| | $b$ | 95CI | $p$ | | $b$ | 95CI | $p$ | |
| (Intercept) | 86.2 | -94.2;266.6 | .343 | | 12.0 | 10.3;13.7 | $< 2 \times 10^{-16}$ | *** |
| Age | -73.3 | -198.6;117.4 | .246 |  | -0.5 | -1.7;0.7 | .412 |  |
| Sex | -44.0 | -215.5;127.5 | .610 |  | -0.2 | -1.8;1.4 | .810 |  |
| Body mass index | -29.1 | -125.1;66.8 | .546 |  | -0.5 | -1.5;0.4 | .237 |  |
| Pain | 13.7 | -74.0;101.4 | .756 |  | 0.01 | -0.8;0.8 | .976 |  |
| Chronic conditions | 24.5 | -42.3;91.3 | .467 |  | -0.3 | -0.9;0.4 | .382 |  |
| Fear of movement | 27.3 | -62.9;117.4 | .547 | | -2.0 | -2.9;-1.2 | $1.2 \times 10^{-5}$ | *** |

Notes: \* $p < .05$ ; \*\* $p < .01$ ; \*\*\* $p < .001$ ; $b$ = regression coefficient; 95CI = 95% confidence interval

#### 3.2.3. Moderation analysis

In the sample of participants with osteoarthritis (n = 68), the moderation analysis showed an interaction effect between fear of movement and approach-avoidance tendencies toward physical activity on usual physical activity levels (b = 134.1; 95CI = 18.0 to 250.2; p = .024) (Moderation 1; Figure 1; Figure 4; Table 3). A simple effects analysis revealed that fear of movement was negatively associated with usual physical activity levels when participants had a tendency to avoid physical activity (i.e., mean neutral-adjusted reaction time to avoid physical activity stimuli – mean neutral-adjusted reaction time to approach this type of stimulus < 0 ms) or a weak tendency to approach physical activity (< 101 ms). However, when the mean neutral-adjusted reaction time required to approach physical activity stimuli was 101 ms or faster than the mean neutral-adjusted reaction time to avoid such stimuli (indicating a stronger tendency to approach physical activity), fear of movement was no longer associated with lower levels of physical activity (Figure 4).

**Figure 4.**
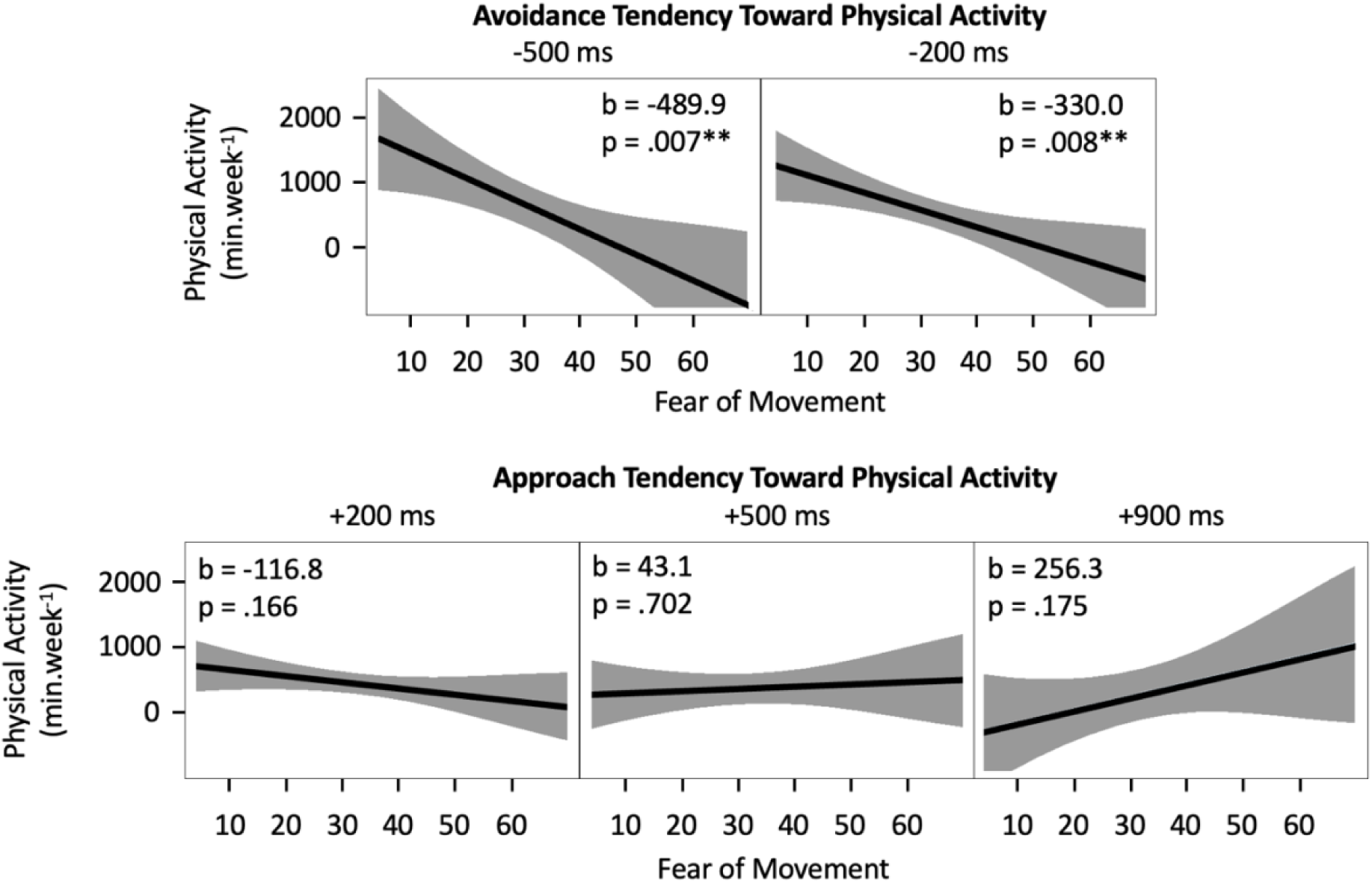
Illustration of Moderation 1: Simple effect of fear of movement on usual physical activity level as a function of approach or avoidance tendency toward physical activity, measured by mean neutral-adjusted reaction time difference (avoid physical activity stimuli – approach physical activity stimuli). A negative difference indicates faster avoidance of physical activity stimuli (i.e., avoidance tendency; top panel), and a positive difference indicates faster approach (i.e., approach tendency; bottom panel). **p<.01

**Table 3.** Moderation analysis: Estimated effects on the usual level of moderate-to-vigorous physical activity (n = 68 participants with osteoarthritis).

| Variables | b | 95CI | $p$ | |
| --- | --- | --- | --- | --- |
| (Intercept) | 474.1 | 182.0;766.1 | .002 | ** |
| Age | -297.1 | -502.3;-92.0 | .005 | ** |
| Sex | 250.3 | -33.7;534.2 | .083 |  |
| Body mass index | 76.2 | -79.1;232.2 | .332 |  |
| Pain | 94.6 | -43.1;232.3 | .174 |  |
| Chronic conditions | 15.6 | -92.7;124.0 | .774 |  |
| Approach-avoidance tendencies | -84.9 | -187.1;17.2 | .101 |  |
| Explicit attitudes | 84.1 | -68.1;236.3 | .273 |  |
| Fear of movement | -177.8 | -349.0;-6.6 | .042 | * |
| Fear of movement × Approach-avoidance tendencies | 134.1 | 18.0;250.2 | .024 | * |
| Fear of movement × Attitudes | 66.1 | -35.0;167.3 | .196 | |

Notes: \* $p < .05$ ; \*\* $p < .01$ ; b = regression coefficient; 95CI = 95% confidence interval

The interaction between fear of movement and explicit attitudes was not significant (b = 66.1; 95CI = -35.0 to 167.3; p = .196) (Moderation 2; Table 3).

## 4. DISCUSSION

### 4.1. Main findings

Our results showed an association between fear of movement and usual moderate-to-vigorous physical activity levels in people with osteoarthritis. In this population, higher fear of movement was associated with lower physical activity engagement, but only in participants with an automatic tendency to avoid physical activity stimuli or a weak tendency to approach them. In addition, contrary to our hypothesis, greater fear of movement was associated with stronger automatic tendencies to approach physical activity and to avoid sedentary behaviors.

### 4.2. Fear of movement and physical activity in people with arthritis

Our results support previous literature showing a negative association between fear of movement and physical activity in people with osteoarthritis.^29–32^ These results suggest that osteoarthritis creates physical and psychological conditions^1^ that make people more susceptible to the effects of fear of movement, thereby moderating its relationship with physical activity. Promoting gradual and supervised physical activity through education and reassurance from a health professional may help to mitigate this moderation effect.^33^ However, this process may be challenging in patients with chronic conditions such as arthritis, where pain is a common symptom that reduces the capacity to associate enjoyment and physical activity. Future work is needed to better understand how intervention targeting this association can be tailored to individuals with chronic conditions.

### 4.3. Moderation by approach-avoidance tendencies

Our results showed that the negative association between fear of movement and physical activity was dependent on the automatic tendency to approach or avoid physical activity stimuli. Specifically, this association became nonsignificant when approach tendencies toward physical activity were stronger. This finding highlights the importance of approach-avoidance tendencies in regulating physical activity behavior and is consistent with intervention studies targeting automatic attitudes. These interventions have been successful in modifying unhealthy behaviors, such as alcohol consumption, smoking, and eating behavior.^34,35^ Whether such interventions can improve physical activity engagement is currently being investigated^36–39^ and could benefit from new robotic tools.^40^

Avoidance tendencies toward physical activity result from the automatic activation of unpleasant affective memories that have been associated with the concept of physical activity during past experiences.^11^ Because our results suggest that avoidance tendencies potentialize and amplify the detrimental effects of fear of movement, health professionals should aim to minimize the risk for such an association in their interventions by fostering pleasurable and emotionally positive physical activity experiences. This suggestion is consistent with existing literature highlighting the role of pleasure and displeasure in physical activity engagement.^41,42^

### 4.4. Limitations

Our results should be considered in light of potential limitations. First, arthritis status was self-reported, which may introduce measurement bias and lead to misclassification of participants. However, the agreement between self-reported arthritis and medical records ranges from 71% for osteoarthritis to 91% for rheumatoid arthritis.^43^ Second, the high self-reported physical activity levels should be considered in light of evidence that the IPAQ-SF overestimates physical activity by an average of 84% when compared to objective measures.^44^ Assessing physical activity and sedentary behaviors using device-based measures would have provided more reliable estimates. Third, the online nature of the study made it impossible to limit the influence of potential distractions in the participant’s environment (e.g., noise that would detract from the task) and to control whether participants were using their two index fingers to perform the task as instructed and whether they were sitting or standing, which may have influenced the results.^45^ Fourth, the format of the modified version of the Tampa Scale of Kinesiophobia, which uses a 7-point scale instead of the 4-point scale, has not been validated. However, statistical properties (e.g., variance, skewness) have been shown to be similar across scale format.^46^ In addition, the items of this modified scale are validated as they are the same as in the original scale. Fifth, the level of arthritis was not assessed, which may influence psychological processes and contribute to heterogeneity within the osteoarthritis group. Future studies should control or test for the potential effect of arthritis severity.

### 4.5. Conclusion

Although fear of movement hinders engagement in physical activity in people with osteoarthritis, interventions that target the underlying mechanisms of approach-avoidance tendencies may overcome this barrier. Physiotherapists and other health professionals should not only promote physical activity,^47^ but also ensure that it is associated with positive emotional experiences. These positive associations may reshape automatic tendencies to counteract avoidance behaviors driven by fear of movement and contribute to increased engagement in physical activity.

## Data Availability

All data produced are available online at https://doi.org/10.5281/zenodo.14715426

## 5. DECLARATIONS

### 5.1. Data and code availability

According to good research practices,^47^ the dataset and the R script are available in Zenodo.^48^ This manuscript was published before peer review on the MedRxiv preprint repository on January 25, 2025.^49^ This preprint version includes additional analyses based on linear mixed effects models testing whether fear of movement was associated with approach-avoidance tendencies. This analysis was not included in the present article due to word limit policy.

### 5.2. Authorship contribution statement

Based on the Contributor Roles Taxonomy (CRediT),^50^ individual author contributions to this work are as follows: Miriam Goubran: Conceptualization; Investigation; Writing – Original Draft; Christian Zammar: Investigation; Santiago Tellez Alvarez: Investigation; Élodie Heran: Investigation; Sara Proulx: Investigation; Martin Bilodeau: Conceptualization; Writing – Original Draft; Supervision (MG); Project Administration; Matthieu P. Boisgontier: Conceptualization (Lead); Methodology; Formal Analysis; Data Curation; Visualization; Writing – Original Draft (Lead); Writing – Review and Editing; Supervision (MG, CZ, STA, EH, SP); Project Administration; Funding Acquisition.

### 5.3. Reporting guidelines

This manuscript conforms to the STROBE guidelines for observational studies.^51^

## 5.6. Acknowledgment

The authors are thankful to the Arthritis Society of Canada for their contribution to the recruitment of the participants. DeepL was used to refine the language and improve readability of this manuscript.

## SUPPLEMENTARY MATERIAL

**Supplementary Table 1.** Estimated effects on the usual level of moderate-to-vigorous physical activity (n = 184 including 68 participants with osteoarthritis and 116 without arthritis).

| Variables | b | 95CI | p |  |
| --- | --- | --- | --- | --- |
| (Intercept) | 470.9 | 254.4;687.4 | $2.9 \times 10^{-5}$ | *** |
| Age | -42.0 | -135.6;51.0 | .373 |  |
| Sex | 196.4 | 34.0;358.9 | .018 | * |
| Body mass index | 35.7 | -43.8;115.1 | .377 |  |
| Pain | -3.3 | -95.3;88.6 | .943 |  |
| Chronic conditions | -22.6 | -90.6;45.3 | .511 |  |
| Osteoarthritis status (presence vs. absence) | -86.7 | -275.8;-13.2 | .366 |  |
| Fear of movement | -150.4 | -287.5;-13.2 | .032 | * |
| Fear of movement × Arthritis status | 193.7 | -23.6;363.8 | .026 | * |

Notes: \*p<.05; \*\*\*p<.001; b = regression coefficient; 95CI = 95% confidence interval

**Supplementary Figure 1.**
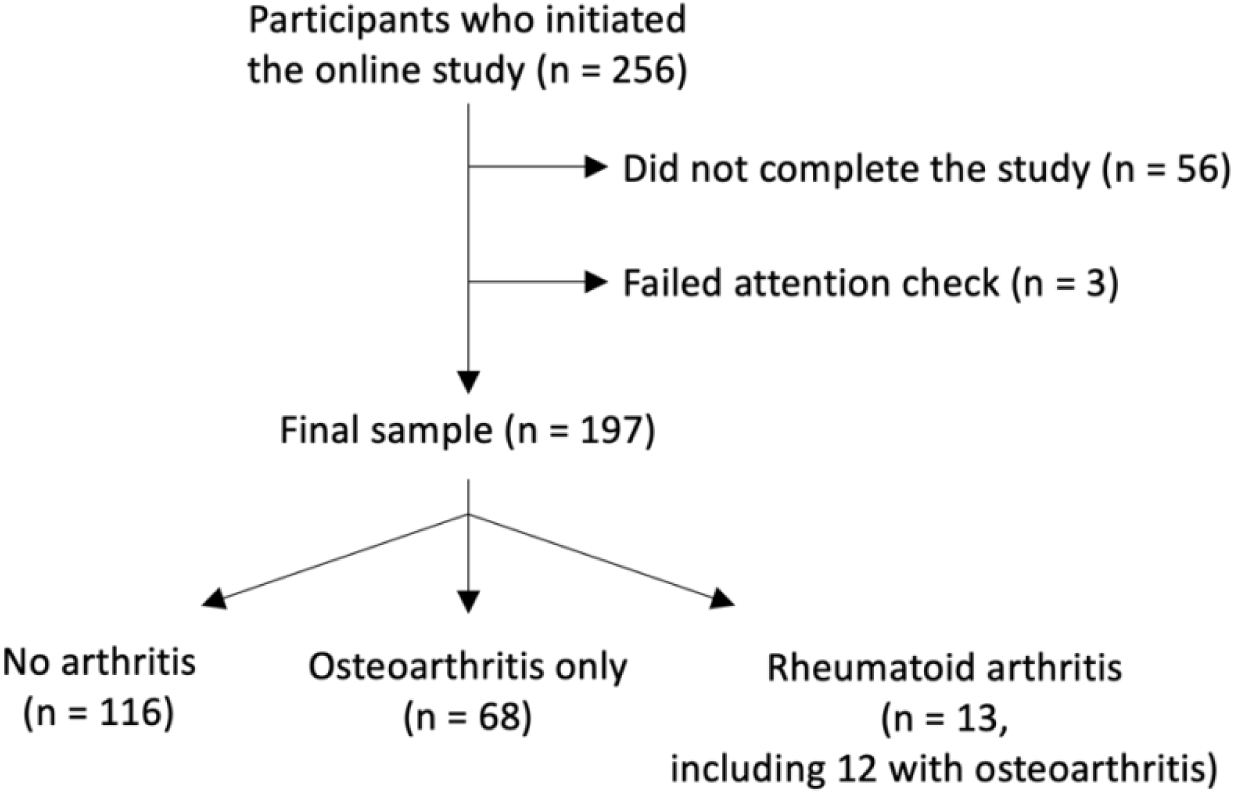
Participant flowchart.

## Supplementary Material 1. Variables

### Chronic conditions

The other possible answers to the question based on item PH006 of the Survey of Health, Ageing and Retirement in Europe were ‘A stroke or cerebral vascular disease’, ‘High blood pressure or hypertension’, ‘High blood cholesterol’, ‘Diabetes or high blood sugar’, ‘Chronic lung disease such as chronic bronchitis or emphysema’, ‘Asthma’, ‘Osteoporosis’, ‘Cancer or malignant tumour, including leukaemia or lymphoma, but excluding minor skin cancers’, ‘Stomach or duodenal ulcer, peptic ulcer’, ‘Parkinson’s disease’, ‘Hip fracture or femoral fracture’, ‘Alzheimer’s disease, dementia, organic brain syndrome, senility or any other serious memory impairment’, ‘Other affective or emotional disorders, including anxiety, nervous or psychiatric problems’, ‘Chronic kidney disease’, ‘Other conditions, not yet mentioned’, and ‘None’.

### Fear of movement

A modified version of the 11-item Tampa Scale of Kinesiophobia (TSK) assessed the participants’ fear of injury during exercise, their susceptibility to injury, the evolution of their pain should they try to overcome it or to exercise, and the safety of physical activity for people with their condition. The instructions to the participants were as follows: ‘Please answer the following questions according to your true feelings, not according to what others think you should believe. Score each statement from strongly disagree (1) to strongly agree (7) by tapping the appropriate box. Select ‘NA’ if the statement is not applicable to you.’ Therefore, the score ranged from 0 (only NAs) to 77. The 7-point scale was used instead of the 4-point scale to avoid forcing the participant to commit to a certain position when they did not have a definite opinion on the statement. The “NA” option was added so that participants could choose not to respond if the statement was not applicable to them (e.g., statements related to pain increase for participants who did not experience any pain).

### Automatic approach-avoidance tendencies

The approach-avoidance task includes two experimental conditions and two neutral conditions (Cheval et al., 2018; Farajzadeh et al., 2023, 2024). In the experimental conditions, each trial begins with a fixation cross displayed in the center of the screen for a random amount of time ranging from 500 to 750 ms (Figure 2A). An avatar then appears in either the upper or lower third of the screen for 1 second, followed by a pictogram in the center of the screen representing an active or sedentary behavior (Figure 2A). The participant, seated in front of the computer with their index fingers positioned on the ‘U’ (top) and ‘N’ (bottom) keys, is instructed that pressing the top key moves the avatar upward and pressing the bottom key moves it downward. The action (approach or avoidance) depends on the avatar’s initial position. When the avatar appears below the stimulus, the top key performs an approach movement, while the ‘N’ key performs an avoidance movement. Conversely, when the avatar appears above the stimulus, the approach and avoidance movements are reversed: The top key performs an avoidance movement and the bottom key performs an approach movement.

In one experimental condition, participants were instructed to quickly move the avatar toward (approach) pictograms depicting physical activity and move the avatar away from (avoid) pictograms depicting sedentary behavior (Figure 2B). In the other experimental condition, the instructions were reversed: Participants moved the avatar away from physical activity and toward sedentary stimuli. The order of the experimental conditions was randomized across participants. The neutral conditions were included to account for generic approach-avoidance tendencies (i.e., not tied to a specific type of stimulus) that might differ across participants (Farajzadeh et al., 2023). In these neutral conditions, the pictograms representing physical activity and sedentary behaviors were replaced with abstract stimuli (rectangles or ellipses) that matched the number and size of information present in three physical activity stimuli (swimming, hiking, cycling) and three sedentary stimuli (couch, hammock, reading). Two neutral conditions were tested. In one condition, participants were instructed to move the avatar toward stimuli with circles and away from stimuli with squares. In the other condition, the instructions were reversed. The order of the neutral conditions was randomized.

One neutral condition was tested before and the other neutral condition was tested after the two experimental conditions. Each condition contained 96 stimuli, 48 of each type (physical activity and sedentary stimuli in the experimental conditions; rectangles and ellipses in the neutral conditions), presented in random order. Task familiarization was performed during the first 15 trials of the study, which were excluded from the analyses. Additionally, the first three trials of each subsequent condition served as familiarization for that condition and were also excluded from the analyses. Before starting each experimental condition, all physical activity and sedentary stimuli were displayed on the screen for seven seconds to ensure participants were familiar with the stimuli. Participants were allowed to rest for as long as needed between conditions, resuming the task by pressing the space bar. During the task, if the participant pressed the incorrect key, the message ‘error’ was displayed on the screen for 800 ms before the next trial. Similarly, if the reaction time, measured as the interval between stimulus appearance and key press, exceeded seven seconds, the message ‘too slow’ was displayed for 800 ms before the next trial (Figure 2A). Automatic approach-avoidance tendencies were derived from reaction times, i.e., the time between stimulus appearance and key press. For each trial in the experimental conditions (i.e., physical activity and sedentary stimuli), reaction time was adjusted by subtracting the mean reaction time for approaching or avoiding neutral stimuli from the raw reaction time of that trial. The tendency to approach physical activity stimuli was computed by subtracting the mean neutral-adjusted reaction time to approach these stimuli from the neutral-adjusted reaction time to avoid them. This subtraction (avoid – approah) resulted in positive values indicating a tendency to approach physical activity (faster approach), whereas negative values indicated a tendency to avoid physical activity (faster avoidance). Incorrect responses, responses faster than 150 ms, and responses slower than 3,000 ms were excluded from the analyses to account for outliers and lapses in attention (Farajzadeh et al., 2023, 2024).

A previous study (Farajzadeh et al., 2023) showed good internal consistency (Spearman-Brown corrected splithalf = 0.83; 95 Confidence Interval = 0.77 – 0.88).

## Supplementary Material 2. Rheumatoid arthritis

To test the effect of fear of movement on usual physical activity levels in people with and without rheumatoid arthritis, we performed a multiple linear regression analysis testing the interaction between fear of movement and arthritis status (rheumatoid arthritis vs. no rheumatoid arthritis) and including five control variables (age, sex, body mass index, pain, and number of chronic conditions).

Thirteen participants with rheumatoid arthritis (age: 55.2 ± 19.5 years, physical activity: 313.8 ± 658.5 min/week, fear of movement: 40.0 ± 15.4, pain: 4.4 ± 1.3, body mass index: 28.4 ± 7.0 kg/m^2^, chronic diseases: 3.8 ± 2.0, 10 female participants, 3 male participants; including 12 participants who also had osteoarthritis) were included in this analysis.

Results showed no evidence of this interaction in people with rheumatoid arthritis (b = -203.8; 95CI = -477.0 to 69.4; p = .143). However, this analysis was not adequately powered. If the sample size is too small to achieve adequate statistical power, variability in the data is high. As a result, any significant or nonsignificant effect may be due to an overestimation or underestimation of the true effect size, respectively (Brysbaert, 2019). In such cases, as for our rheumatoid arthritis subgroup, the results should be considered exploratory and should be confirmed in subsequent studies with adequate power.

